# A systematic review of antibody mediated immunity to coronaviruses: antibody kinetics, correlates of protection, and association of antibody responses with severity of disease

**DOI:** 10.1101/2020.04.14.20065771

**Authors:** Angkana T. Huang, Bernardo Garcia-Carreras, Matt D.T. Hitchings, Bingyi Yang, Leah C. Katzelnick, Susan M. Rattigan, Brooke A. Borgert, Carlos A. Moreno, Benjamin D. Solomon, Isabel Rodriguez-Barraquer, Justin Lessler, Henrik Salje, Donald Burke, Amy Wesolowski, Derek A.T. Cummings

## Abstract

The duration and nature of immunity generated in response to SARS-CoV-2 infection is unknown. Many public health responses and modeled scenarios for COVID-19 outbreaks caused by SARS-CoV-2 assume that infection results in an immune response that protects individuals from future infections or illness for some amount of time. The timescale of protection is a critical determinant of the future impact of the pathogen. The presence or absence of protective immunity due to infection or vaccination (when available) will affect future transmission and illness severity. The dynamics of immunity and nature of protection are relevant to discussions surrounding therapeutic use of convalescent sera as well as efforts to identify individuals with protective immunity. Here, we review the scientific literature on antibody immunity to coronaviruses, including SARS-CoV-2 as well as the related SARS-CoV-1, MERS-CoV and human endemic coronaviruses (HCoVs). We reviewed 1281 abstracts and identified 322 manuscripts relevant to 5 areas of focus: 1) antibody kinetics, 2) correlates of protection, 3) immunopathogenesis, 4) antigenic diversity and cross-reactivity, and 5) population seroprevalence. While studies of SARS-CoV-2 are necessary to determine immune responses to it, evidence from other coronaviruses can provide clues and guide future research.

**Key Questions:** **Key Questions for SARS-CoV-2**

- What are the kinetics of immune responses to infection?
- Do people who have more severe disease mount stronger antibody responses after infection?
- How do antibody responses vary between different types of antibodies or as measured by different assays?
- How does the presence of antibodies impact the clinical course and severity of the disease?
- Is there cross-reactivity with different coronaviruses?
- Does cross-reactivity lead to cross-protection?
- Will infection protect you from future infection?
- How long will immunity last?
- What are correlates of protection?

## Introduction

A pandemic of severe acute respiratory syndrome coronavirus 2 (SARS-CoV-2) is currently underway resulting in worldwide severe morbidity and mortality. Limited pre-existing immunity to this virus is thought to be responsible for the explosive increase in cases across the world. Nearly all transmission models of SARS-CoV-2 assume that infection produces immunity to reinfection for durations of at least one year^1–3^. This assumption is relevant to public health officials implementing and managing various non-pharmaceutical interventions, the utility of sera from infected individuals as a therapeutic^4^, and the ability for serological tests to identify those who are immune^5^. The dynamics of immunity will also affect the performance of serological testing to quantify the extent of infection in populations. However, knowledge of the dynamics and nature of immune response to SARS-CoV-2 infection is limited, and the scientific basis for durable immunity, upon which these key public health and clinical strategies are dependent, is not well developed.

Several authors have noted human experimental infection studies (called human challenge studies) suggesting that protection after coronavirus infections may last only 1 or 2 years^6–9^. HCoVs have been used in human challenge experiments since shortly after their discovery in 1965^10,11^. These experiments, where individuals were intentionally infected with HCoV, provide some of the clearest characterization of human responses to coronaviruses and the potential for immune responses to limit infection and disease. Multiple human challenge studies measured antibody immunity before a coronavirus challenge and identified antibody responses that were associated with protection from infection, serological response or symptom^12^. The low severity of HCoV allowed for safe use of these viruses in human challenge experiments. The greater likelihood of severe illness in SARS-CoV-2 limits the applicability of such experiments, although some have argued for their use in subsets of the population^13^.

The duration of immunity of SARS-CoV-2 will dictate the overall course of the pandemic and the post-pandemic dynamics^7^, and so an understanding of the temporal dynamics of protective immunity is critical. As with other introductions of novel pathogens^14^, explosive outbreaks of SARS-CoV-2 across the globe may threaten its persistence by reducing the number of available hosts susceptible to infection. Immune interactions with endemic coronaviruses could theoretically affect the short- and long-term dynamics of SARS-CoV-2, and vice versa^15,16^ through cross-protection or antibody-dependent enhancement^17^, but these interactions and effects are not yet understood. If SARS-CoV-2 does become endemic, age-stratified seroprevalence studies of endemic coronaviruses may provide estimates for incidence rates in the presence of higher levels of population immunity.

Here, we describe the results of a systematic review of the literature on antibody measures of immunity to coronaviruses, including endemic human coronaviruses (principally HCoV-229E, HCoV-HKU1, HCoV-OC43, HCoV-NL63), SARS-CoV-1, MERS-CoV (Middle East respiratory syndrome CoV) and early work on SARS-CoV-2. We conceptualize the stages of exposure and infection at which immunity may play a role in the dynamics of SARS-CoV-2, and how literature describing work on this and other coronaviruses can provide insights into these stages, as follows (see Figure 1, a visual abstract of our review). First, an exposure to a pathogen generates an immune response that changes over time and between individuals (**antibody kinetics**). Upon exposure, infection history might play a role in providing protection against new infection, and the literature can provide evidence for such **correlates of protection** through challenge studies and longitudinal cohort studies. Upon infection, an individual’s immune state, possibly impacted by pre-existing antibodies to other coronaviruses (among other mechanisms), may cause harm through **immunopathogenesis**, and the literature on this topic primarily consists of *in vitro* experiments. Correlates of immunity (or risk through immunopathogenesis) are complicated by the existence of multiple genera of coronaviruses that are **antigenically diverse** and may provide cross-protection, and may also cause false positive assay results due to **cross-reactivity**. Finally, the preceding phenomena at the individual level interact to determine **population seroprevalence**. Studies that measure these quantities across different age groups can provide evidence, albeit ecological evidence, for or against proposed mechanisms of immunity at the individual level.

**Figure 1:**
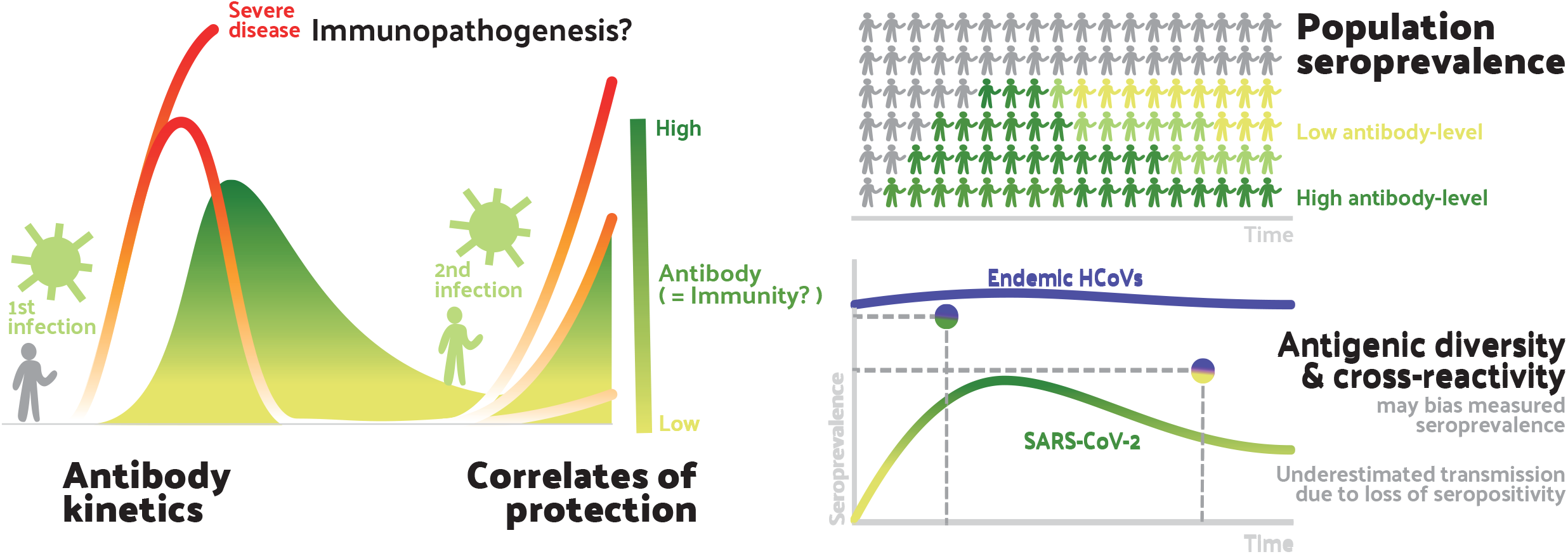
Aspects of antibody response included in this review. This figure shows the areas of focus of our review within our conceptualisation of the stages of exposure and infection at which we believe antibody mediated immunity may play a role in the dynamics of SARS-CoV-2. At the individual level (left), antibody response following the first infection/exposure increases and then declines (**Antibody kinetics**). Sometime later individuals may be exposed to SARS-CoV-2 again. They may be protected from infection by their acquired immunity (**Correlates of protection**). Their acquired immunity may also moderate the severity of infection with some possibility that pre-existing immunity may lead to **immunopathogenesis** (relevant to both first and second exposure). These individual-level dynamics aggregate to form the **population-level seroprevalence** (right-top). Measures of seroprevalence may imperfectly measure past exposure to infection due to **antigenic diversity** of future SARS-CoV-2 viruses and **cross-reactivity** of endemic human coronaviruses (HCoVs) with SARS-CoV-2. Measures of seroprevalence may also be inconsistent across times as antibodies-levels within individuals wane.

## Methods

### Search strategy and selection criteria

We conducted searches of the PubMed database on March 20, 2020 using the search term “coronavirus” and each of the following terms or phrases: “serolog*”, “serop*”, “cross reactivity”, and “complement fixation”. We also searched for each of these terms with the search terms “SARS” and “MERS”. Articles were included in the search regardless of publication date. Articles included electronic, ahead-of-print publications available in the PubMed database. We did not attempt to contact authors to obtain unpublished data. Each abstract was reviewed by two reviewers. Articles that were not considered applicable were excluded (see below). Additionally, we identified articles that may be relevant from the reference list of other articles identified by our PubMed searches.

### Assessment

Abstracts were classified as: 1) study of antibody immune responses to human coronavirus; 2) study of human coronavirus but not about antibody immunity; 3) study in non-humans or basic biological study of virus-antibody interactions not characterizing actual humans; 4) study in humans not about coronavirus; 5) in another language; 6) duplicate of another record; or 7) dead link/bad return/noise. Disagreement between two reviewers on classification was resolved by a third reviewer. Manuscripts classified in the first category (study of antibody immune responses to human coronavirus) were reviewed in their entirety and classified by one reviewer as relevant to one or more of the following: 1) antibody kinetics; 2) correlates of protection; 3) antigenic diversity and cross reactivity; 4) immunopathogenesis; and/or 5) population seroprevalence or incidence. Manuscripts that were not considered relevant to any of the preceding categories were excluded.

### Data Abstraction

Data were digitized from papers according to pre-defined criteria for each area of focus, related to accessibility, utility in answering the key questions and suitability for the pooled analyses (see Supplementary Appendix for details). Data are available at https://doi.org/10.5281/zenodo.3751566.

### Pooled Analyses

We conducted pooled analyses in two areas where data were available and sufficiently comparable to include in a single analysis.

#### Antibody kinetics and association of antibody responses with clinical severity

We summarized approximate time to detection across studies for two sets of studies. The first set includes studies with cumulative number of positive cases over time. Each additional positive case added to the cumulative curve was assumed to be a new detected case at that time point. This is approximate only, because cumulative curves sometimes decline (Figure S1), indicating some participants were lost to follow-up. Second, we identified data that described either seropositivity as a function of time, or measures of antibodies with a reported cutoff. Data was included if (i) the first seropositive time point was within three weeks of illness onset or (ii) the first estimate(s) were negative, but a subsequent measurement within three weeks of the first was positive.

#### Population seroprevalence

We estimated the annual force of infection from age-stratified seroprevalence data, assuming that individuals acquire infection with a single, strain-specific force of infection that is constant in time using standard methods^18^, and that once an individual becomes seropositive they remain seropositive for the remainder of their lifetime (regardless of their protective status). We estimated the total force of infection (FOI) from the four major endemic strains by drawing 100,000 bootstrap samples of four strain-specific FOIs and calculating the sum across strains. Age at first infection is given by the inverse of total FOI.

## Results

### Manuscript identification

Our searches identified 1281 abstracts of potential relevance (Figure 2). Two reviewers read each abstract and identified 322 manuscripts for full review. Manuscripts were classified into our areas of focus. Below, we present the findings of our review for each area of focus.

**Figure 2:**
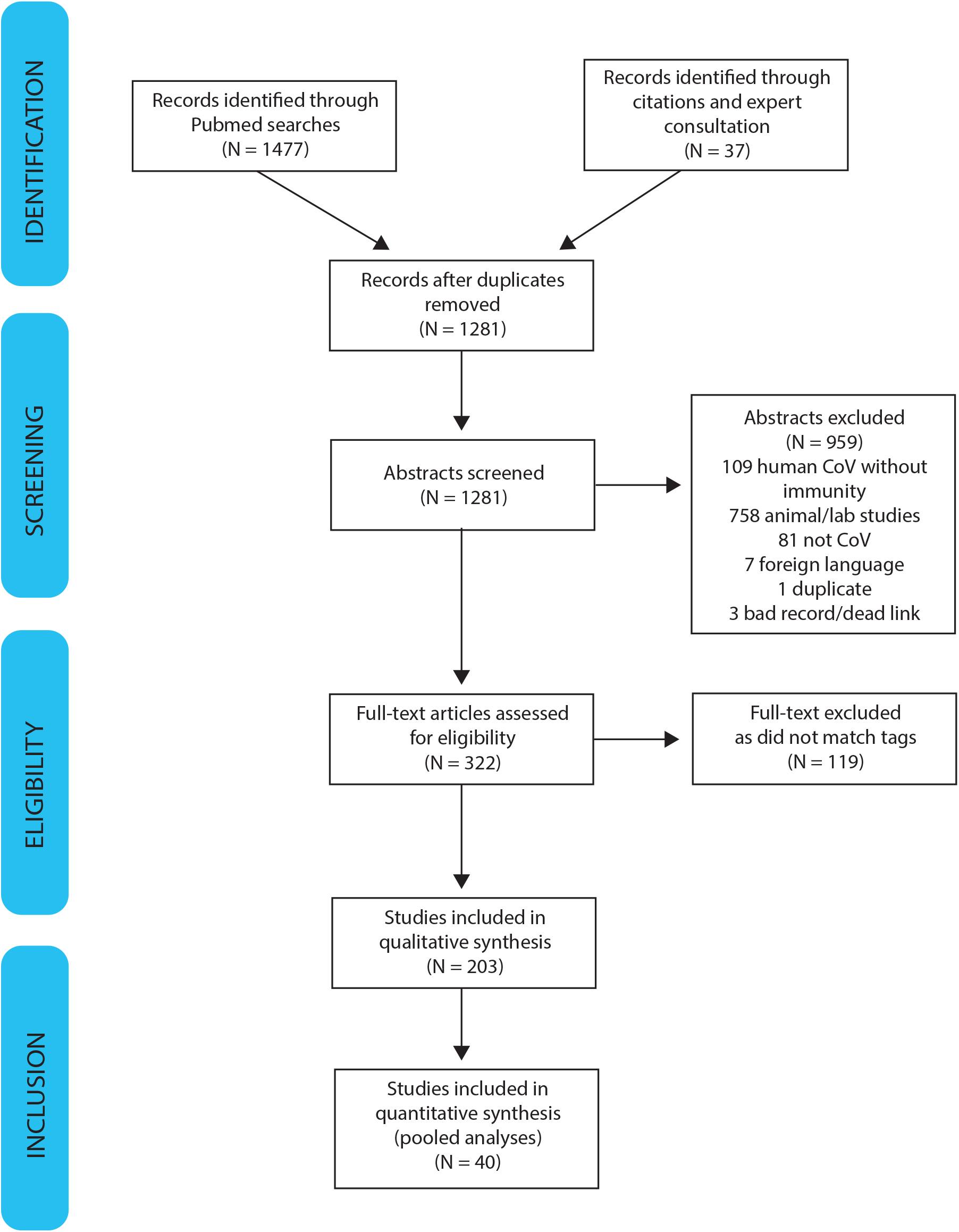
PRISMA diagram of systematic review process.

#### General background for multiple areas of focus: Serological assays

Multiple serological assays have been used to characterize antibody responses to coronaviruses. Assays we encountered in our review fell into two major categories (Table S1). The most commonly used assays were binding assays, including enzyme linked immunosorbent assays (ELISA), immunofluorescence assays (IFA), Western blots, hemagglutination inhibition (HAI), and complement fixation (CF). The second category of assays was neutralization assays. Neutralization assays are typically considered the gold standard for measuring functional antibody responses as they measure biological activity throughout the viral replication process. Researchers have used assays of both types to characterize antibody activity by antibody class (i.e. IgG, IgM, IgA). As these antibodies are known to have different temporal dynamics, we reported when specific classes of antibodies were characterized in figures. If no characterization was specified, we reported the measures of antibody aggregated across all classes. The source of samples in the reviewed studies was almost exclusively serum samples. However, mucosal samples collected by swab or nasal washings were also reported.

### Antibody kinetics and association of antibody responses with clinical severity

Initially, 56 studies were classified as relevant to antibody kinetics, and 40 as relevant to the association of antibody responses with clinical severity. Of these, 27 were selected after further review (4 on endemic HCoV, 10 on MERS-CoV, 11 on SARS-CoV-1, and 3, including 2 non-peer-reviewed preprints, on SARS-CoV-2). Table S2 provides summaries of these studies. We digitized data from a subset of 22 studies that included sufficient detail on longitudinal antibody measurements (Supplementary Data File S1). 18% of studies reported cumulative seroconversion only. 73% of the digitized studies provided estimates during the first week after onset of symptoms, while 77% had measurements at least one month after onset of symptoms.

#### Antibody kinetics post infection

In the studies we shortlisted, antibody responses to infecting coronaviruses were rarely reported during the acute phase of illness (1-7 days)^19–22^. Many studies reported an immune response, characterized by a robust increment of antibody titers for HCoV-229E, MERS-CoV, SARS-CoV-1, and SARS-CoV-2 after the second or third weeks following the onset of illness^8,23–30^.

Callow et al.^8^ found similar dynamics in IgA and IgG across 10 individuals experimentally infected with HCoV-229E: antibody levels increased after 8 days and peaked around 14 days, although significant variation between patients was reported. Yang et al.^31^, analyzing data across 67 patients, found higher positive rates of IgM against SARS-CoV-1 than IgG during the first month. They further found that the proportion of patients who seroconverted for IgM peaked 30 days after onset, followed by a gradual decrease of IgM levels, while IgG levels peaked by week 25^31^. Another study on 30 SARS-CoV-1 infected patients^32^ detected seroconversion of IgG, IgM, and IgA at similar times, indicating that the earliest they reached a peak was on average 15 days. While it is currently too early to characterize how anti-SARS-CoV-2 antibodies will change over prolonged periods of time, preliminary studies have analyzed antibody changes in recent infections. Tan et al.^33^ found that IgM was detected on day 7 and peaked on day 28 (across 28 patients), and IgG appeared by day 10 and peaked on day 49 (45 patients), while Zhao et al.^34^ determined that seroconversion among 173 patients took place at median times of 12 (IgM), 14 (IgG), and 11 (neutralizing antibodies) days.

Multiple studies reported that while IgM and IgG titers increased during the first weeks following symptom onset, IgM levels gradually waned (while remaining detectable) for SARS-CoV-1 and MERS-CoV in comparison to IgG levels about a month post follow-up^23,32^. Most studies that examined antibody kinetics over extended periods of time focused on IgG^32,35–40^. Callow et al.^8^ found among experimental infections with HCoV-229E that after peaks in IgG and IgA, antibody levels waned and between 11 weeks and 1 year post inoculation were at similar levels to those found in inoculated but uninfected patients. Other studies^35,40^ reported detectable IgG levels in recovered MERS-CoV patients, respectively at five months and one year after illness onset, while another^31^ detected IgG antibodies across 67 SARS-CoV-1 patients after 82 weeks, the endpoint of the study. We found few studies that analyzed changes in antibody kinetics over the course of many years after illness onset^36,37^. Cao et al.^37^ described similar long-term dynamics in IgG and neutralizing antibodies over the course of a 3-year study on SARS-CoV-1: titers for both peaked at month 4, and while they waned thereafter, 74.2% and 83.9% of patients had detectable levels of IgG and neutralizing antibodies, respectively, at month 36.

#### Correlation between severe disease and rise in post-infection antibody level

Studies have increasingly explored a potential link between case severity and antibody response; however, the available information analyzing the nature of this relationship is uneven across viruses. Several studies on MERS-CoV as well as the first preliminary analyses on SARS-CoV-2 explicitly explore this connection, while fewer studies on endemic HCoVs and SARS CoV-1 do so. Several studies found that cases of varying severity (e.g., asymptomatic, mild, and severe) developed detectable antibodies against HCoV-229E, SARS-CoV-1, SARS-CoV-2, and MERS-CoV^33,41–44^. One study^45^ found that the rise in antibodies between acute and convalescent sera correlated positively with symptoms and clinical score in 15 patients experimentally infected with HCoV-229E (Table 1). However, most studies of SARS-CoV-1 did not report symptom severity, and evidence of differences in antibody responses among cases experiencing symptoms of different severity is inconclusive. SARS-CoV-1 survivors with sequelae were found to have lower neutralizing antibodies than patients without sequelae^37^, although the same study otherwise found no significant differences in kinetics according to disease severity. Another study^39^ found no evidence of difference in antibody responses between patients who survived or died. In MERS-CoV, on the other hand, Ko et al.^46^ found that both seroconversion rate and peak antibody levels increased with disease severity, while Okba et al.^47^ reported a robust response to severe infections as opposed to low or no seroconversion in asymptomatic and mild cases. Milder infections also appeared to be less likely to elicit serologic responses^47,48^, although Okba et al. suggests that seroconversion detection may depend on the antibodies being targeted. More severe cases were also found to have slower responses^24,27^; 75% of patients who died had not seroconverted by week 3^46^. Some authors have hypothesized^47,49^ that seroconversion rates in severe cases may be associated with prolonged viral shedding, and that low antibody responses in mild cases may be due to short-lived infections. Another study^40^ suggested that weaker antibody responses to endemic HCoVs (specifically HCoV-229E) might be because these mainly infect the upper respiratory tract. Preliminary studies on SARS-CoV-2 point to a possibly contrasting pattern to MERS-CoV: while IgM antibodies appear at the same time in severe and non-severe cases, IgG appears sooner in severe cases^33^. On the other hand, neutralizing antibody titers were higher in severe cases^34^.

**Table 1:**
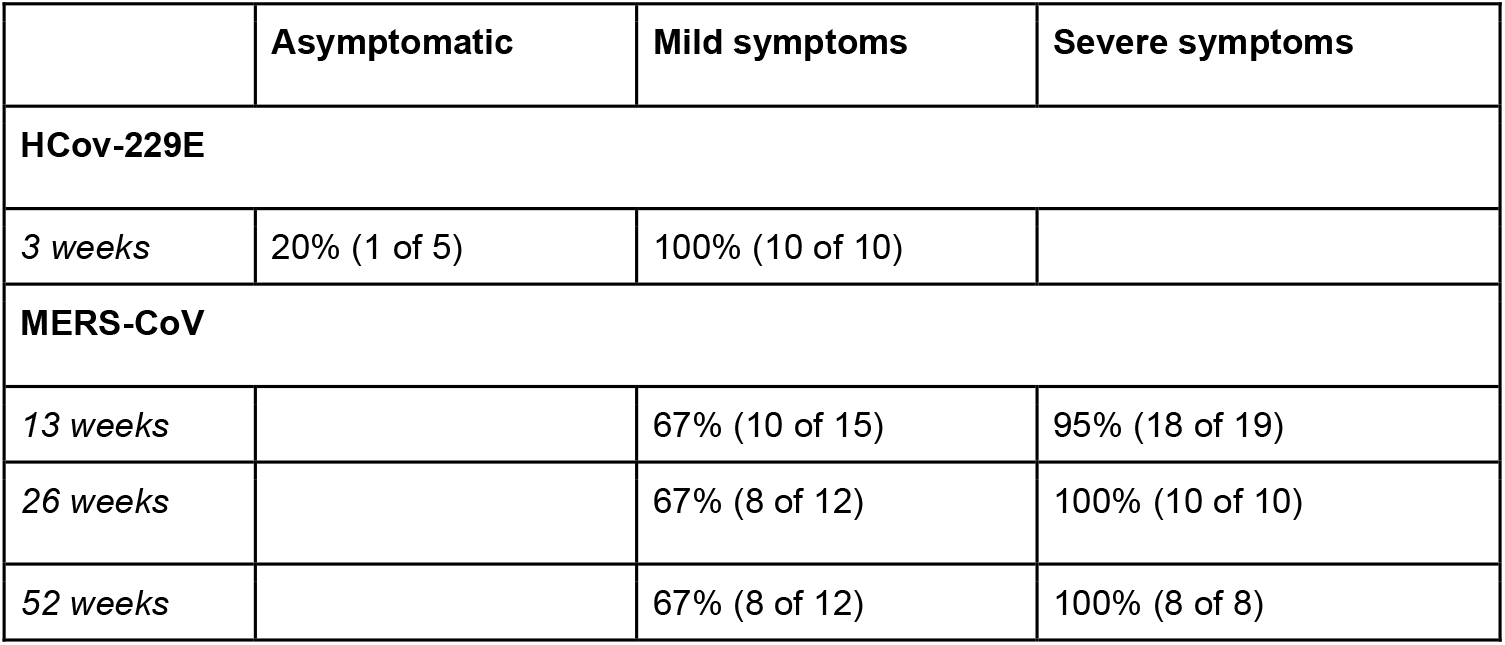
Proportion of patients that had detectable antibodies, at different time points post symptom onset or infection, for HCoV-229E and MERS-CoV. Results for HCoV-229E are from Kraaijeveld et al.^45^, and report the number of individuals experiencing symptoms of a given severity that had significant antibody rises post infection. Results for MERS-CoV are taken across studies in which a cutoff for the assay being used was provided, and gives the number of patients for which IgG levels were above the respective cutoffs, at different time points (also see Figure 4). For example, at week 26, there were 12 patients with mild symptoms from studies that reported a cutoff, and 8 of those patients had IgG levels above that cutoff. MERS-CoV data were digitised from six studies ^35,40,46,47,142,143^

The distributions of time points at which antibodies were detected (see Methods) in the digitized data are shown in Figure 3. The median time to detection was similar across different antibodies for SARS-CoV-1 (12 days; IQR 8-15.2 days) and SARS-CoV-2 (11 days; IQR 7.25-14 days), but longer for MERS-CoV (16 days; IQR 13-19 days). Severity appears to be associated with time to detection of IgM in MERS-CoV cases only (2 days longer), and IgG in both MERS-CoV and SARS-CoV-2 (2 to 3 days longer for more severe cases). All data on time to seroconversion was based on symptomatic patients. No data were available for asymptomatic individuals.

**Figure 3:**
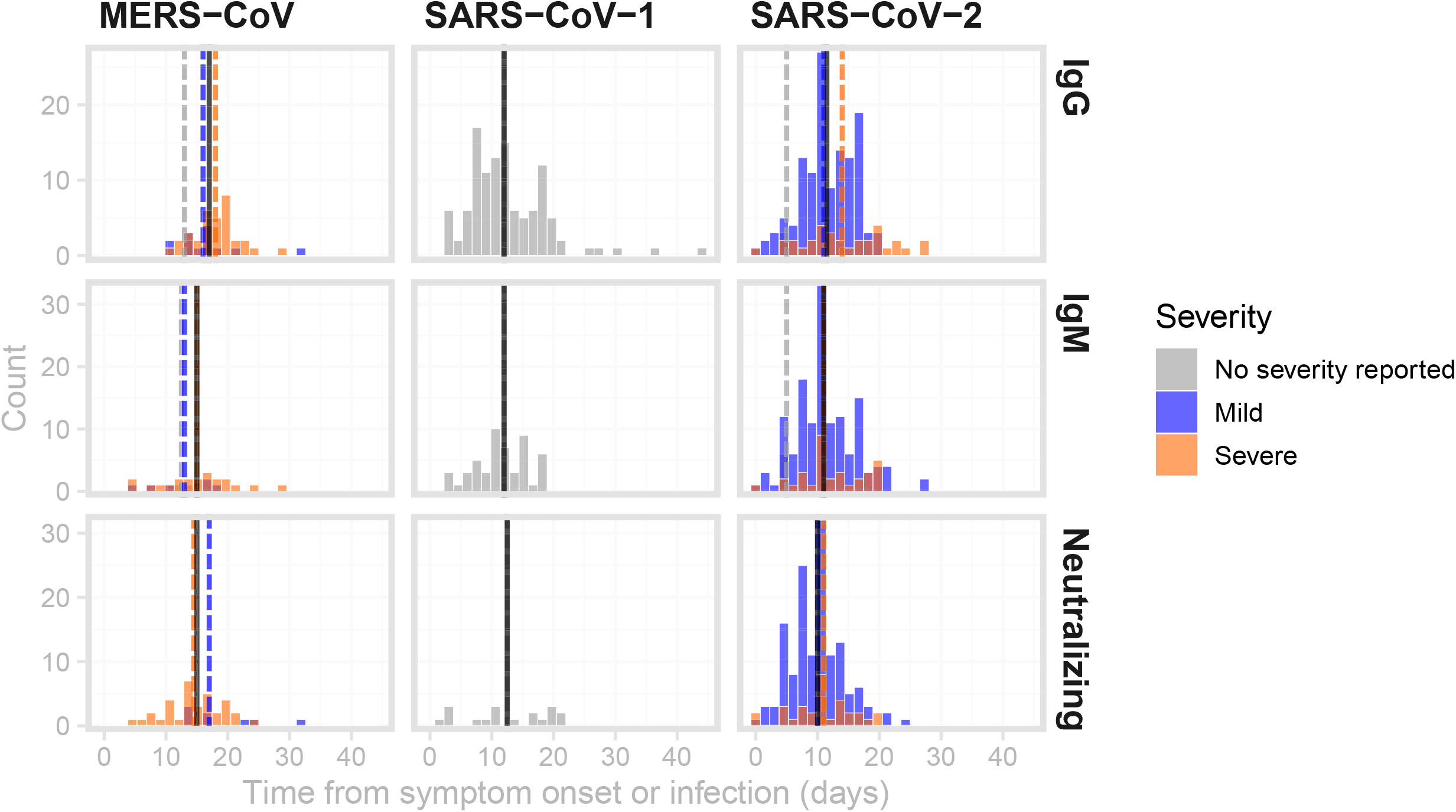
Distributions of times from symptom onset to detection of antibodies against MERS, SARS-CoV-1 and SARS-CoV-2. Times between symptom onset and the detection of IgG (top row), IgM (middle row) and neutralizing antibodies (bottom row). Black vertical lines indicate the median values across all severity ratings, while dashed coloured lines are the median values for mild (blue), severe (orange) illnesses as well as illnesses of unreported severity (grey). Data were digitized from 12 studies^23,27,29,30,32,34,40,44,135–138^.

Figure 4 provides a broad sense for trajectories of antibodies (also see Figures S2 and S3). Most longer-term studies (>10 weeks) were for MERS-CoV and reported IgG and neutralizing antibodies; these showed the presence of IgG and neutralizing antibodies up to 60 weeks after symptom onset (Figure 4). The studies reporting symptom severity with longer-term data focused on MERS-CoV. Not all of these studies reported a cutoff for the assay used; in those that did, two thirds of patients with mild symptoms had detectable or positive IgG antibodies at six months and one year, while all patients with severe symptoms had detectable IgG antibodies at the same points in time (Table 1).

**Figure 4:**
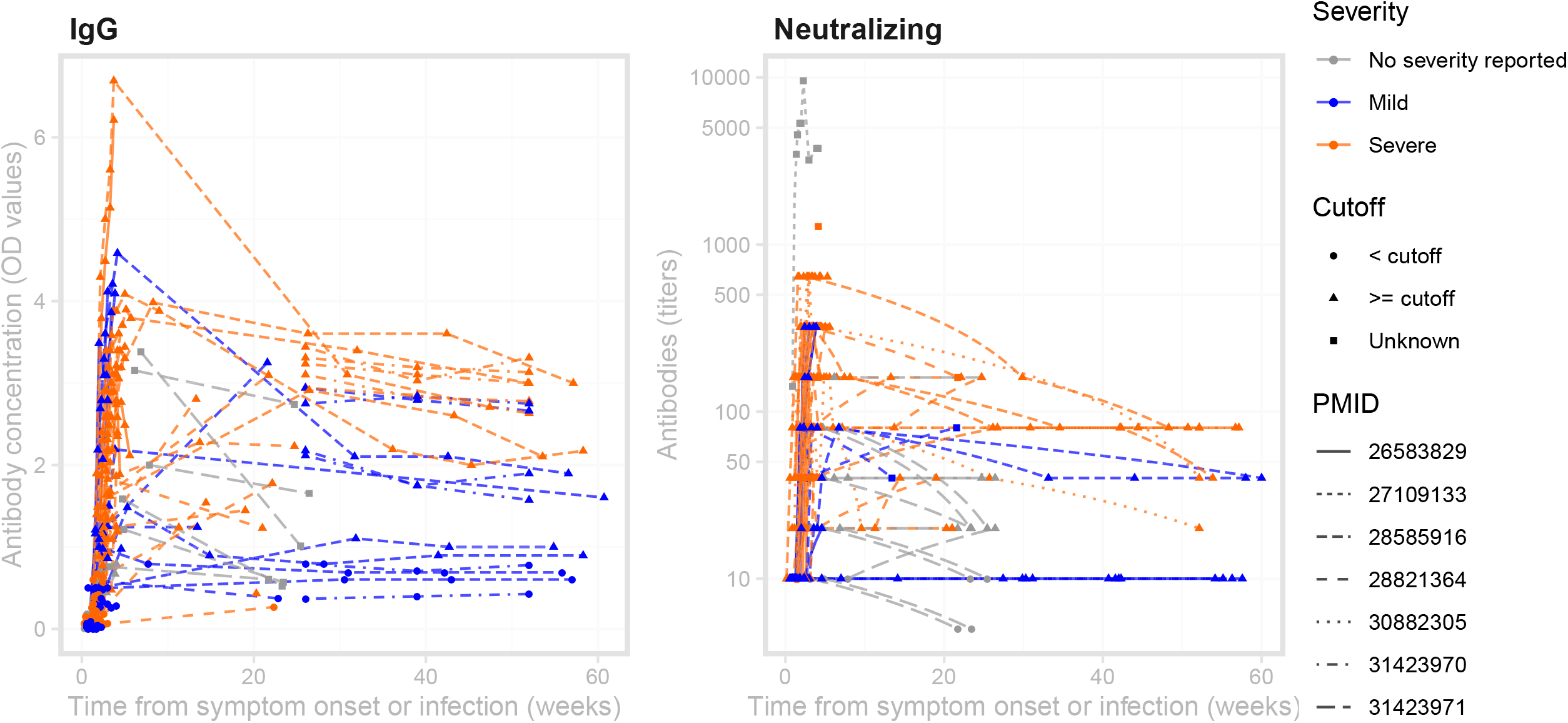
MERS-CoV antibody kinetics. The left panel shows data for studies reporting IgG concentration in units of optical density, while the right panel shows data for studies reporting neutralizing antibodies in units of titers. The plotting symbols indicate whether a measurement was above the cutoff for the assay being used, if reported in the study. Some studies reported titers that were lower than or greater than some threshold value; those are here plotted at those values (e.g., for ≥ 320, the value is assumed to be 320). Some studies may report kinetics of different antibodies or using different assays (and different units) for the same patient. Note that while these are plotted on the same axes, values may not necessarily be comparable across studies within each panel, as each lab may have different assay conditions resulting in different scales. See Figures S2 and S3 for (more limited) data on SARS-CoV-1 and IgA.

### Correlates of protection

Identification of a correlate of protection requires characterization of immune responses prior to a known exposure or period of risk in which infection or illness outcomes are characterized. In our review, we found that this level of detail was only present in human challenge experiments with HCoVs. We identified 18 studies in which volunteers were exposed to experimental infections with HCoV. Of these, 6 associated pre-infection antibody measurements with either virologic, serologic or illness outcomes upon experimental infection (Table S4).

The earliest identified experimental infection study of coronaviruses found that 7 of 8 with neutralizing titer < 5 excreted virus after experimental exposure compared to only 1 of 4 with pre-exposure titer of 40 or greater^10^. Interestingly, this study is one of the few to report illness as a function of dose of viral inoculum given in the challenge experiment and suggests that those given higher doses (>10^1.2^ TCD50) were more likely to experience colds (10/15) than those exposed to lower doses (<10^0.7^ TCD50) (3/11).

Barrow et al.^50^ found lower proportions of individuals with high neutralizing titer experienced ‘significant colds’ upon viral challenge than individuals with low titer.

Callow^12^ characterized IgA, IgG and neutralizing antibody in sera and nasal washings from 33 volunteers before they were experimentally exposed to 229E HCoV. She found that multiple antibody responses were associated with reduced risk of infection, seroconversion, and symptomatic illness upon challenge. Individuals who seroconverted to the experimental viral exposure (defined as a rise in ELISA IgG serum antibodies) had significantly higher serum IgG, neutralizing antibodies, and nasal IgA. Serum and mucosal IgA were associated with the duration of viral shedding post experimental infection, with those shedding for 5 days or more having statistically significantly less mucosal IgA than those shedding less than 5 days (0.6 ng/ml versus 4.7 ng/ml, p<0.01). Serum neutralizing antibody was not found to be significantly associated with viral shedding duration. This study also showed protective associations of pre-infection serum neutralizing antibody, serum IgG and nasal IgA with clinical severity scores and nasal secretion weights (a measure of severity of rhinorrhea symptoms).

Another prospective study^51^ reported detection of pre-existing neutralizing antibodies among medical students who had virus isolation (67%, n=8/12) or seroconversion to HCoV-229E (25%, n=3/12). Pre-existing neutralizing antibodies were inversely associated with increases in neutralizing antibodies after re-infection, but were not associated with reinfection events that were determined by CF seroconversion.

#### Experimental re-infection or re-challenge

Several studies exposed volunteers to two challenges of virus, some months apart. Reed re-challenged 6 volunteers who had been experimentally infected 8-12 months previously^9^. On the first challenge, all 6 developed symptoms and detectable viruses and 5 of 6 experienced significant rise in titer. In the second, 0/6 experienced illness, detectable virus or significant rise in titer. Callow et al re-challenged volunteers with the same dose of coronavirus, one year apart^8^. Of 9 volunteers who were infected in the first exposure, 6 (67%) were infected in the second exposure. However, none of these individuals developed respiratory illness symptoms and they experienced a mean duration of detectable virus of 2 days compared to a mean of 5.6 in the initial challenge.

### Cross-reactivity and antigenic diversity

We identified 73 papers as related to cross-reactivity and/or antigenic diversity (Table S5). Of these studies, 43 were identified as highly important and were described in the text, and data was digitized from 7 studies (Supplementary Data File S2).

#### General background for this area of focus: Genetic relationships of human coronaviruses and immunogenic proteins

Within the *Coronaviridae* family, the *Coronavirinae* subfamily includes four distinct genera. The **Alphacoronaviruses** include two major human coronaviruses, HCoV-229E and HCoV-NL63. Multiple HCoV-229E-like strains have also been characterized. The **Betacoronaviruses** are categorized into 4 lineages. Lineage A includes HCoV-OC43 and HCoV-HKU1, Lineage B includes SARS-CoV-1 and SARS-CoV-2, Lineage C includes MERS-CoV and multiple bat coronaviruses, and lineage D contains coronaviruses thus far only identified in bats. HCoV-OC43 and HCoV-229E are documented to cause common cold while the more recent strains (HCoV-HKU1 and HCoV-NL63) infect both the upper and lower respiratory tract, resulting in more severe but rarely fatal disease^52^. Other CoVs have been associated with human disease, including enteric disease in infants and zoonotic infections from livestock, but seem rare and are not described here^9,53–58^.

Coronaviruses have four structural proteins: the spike protein (S), the nucleocapsid (N), the envelope protein (E) and the membrane protein (M)^59,60^. The S protein, which protrudes from the virus envelope, is immunodominant and consists of two subunits: the S1 protein, which contains the receptor binding domain (RBD) and the S2 protein, which mediates cell membrane fusion^61,62^. The nucleocapsid protein, which is also immunogenic, is smaller than S, lacks a glycosylation site, and induces antibodies sooner than to S during infection, making it an attractive protein for diagnostic assay design^63^. Sequence homology for the N and S of SARS-CoV-1 to other Betacoronaviruses is 33-47% and 29%, respectively, while homology to Alphacoronaviruses is lower (25-29% homology to N and 23-25% for S)^63^. SARS-CoV-2 is most similar to SARS-CoV-1, harboring sequence homology of 90% in N and 76% in S followed by MERS-CoV (48% and 35%, respectively)^64^.

#### Natural and experimental infection studies in humans point to cross-reactivity within but minimal reactivity between endemic human Alpha- and Betacoronaviruses

Individuals experimentally inoculated with HCoV-229E and HCoV-229E-like strain LP experienced a >4-fold rise in neutralizing antibodies to both HCoV-229E and LP, while individuals inoculated with HCoV-OC43 did not^65^. However, while volunteers experimentally inoculated with HCoV-229E-like strains were protected against challenge with the homologous strain at one year (n=6/6; none shed virus, showed symptoms, or had a rise in antibodies), volunteers experienced only partial protection against heterologous HCoV-229E-like strains (n=5/12 protected). Increasing population immunity to HCoV-229E in the population was associated with less clinical disease upon challenge with HCoV-229E-like strains^9^. Studies of serological responses to HCoV N proteins point to cross-reactivity within Alpha-HCoVs (229E and NL63) and Beta-HCoVs (OC43 and HKU1) but not between Alpha-and Beta-HCoVs^66–68^. Consistent with observations in human challenge studies, children experiencing natural HCoV infections experience 4-fold seroconversions to either HCoV-OC43 or HCoV-229E but not both simultaneously^69^. However, a longitudinal study in newborns found that children seroconverted to either HCoV-NL63 or HCoV-229E but not both, although both viruses are Alpha-HCoVs^70^. A later study in newborns followed from age 0 to 20 months^71^ showed asymmetric interactions within Alpha-HCoVs and Beta-HCoVs: seroconversion to HCoV-NL63 was observed after HCoV-229E but no recent HCoV-229E seroconversions had a recent infection by HCoV-NL63, suggesting HCoV-NL63 provides at least short-term protective immunity against HCoV-229E. Similarly, HCoV-HKU1 seroconversion occurred prior to HCoV-OC43 but rarely after, suggesting HCoV-OC43 protected against HCoV-HKU1.

#### Infection with endemic HCoVs produces little cross-reactivity to emerging CoVs SARS-CoV-1 and MERS-CoV

Individuals experiencing natural infections with HCoV-OC43 or HCoV-229E did not have detectable antibodies in acute or convalescent samples against SARS-CoV-1 (to N protein^72^; by IFA or neutralization^39^). Healthy individuals with antibodies to HCoV-229E, HCoV-OC43, and other endemic HCoVs rarely had detectable antibodies that bound SARS-CoV-1 infected cells or SARS-CoV-1 N protein^68,73,74^. HCoV-OC43 N and SARS-CoV-1 N proteins have a subset of sites with shared homology, potentially explaining low-level false positive results for N-based assays^75^. Blood donors in Southern China (n=152) and Saudi Arabia (n=130) did not have detectable binding (IFA) or neutralizing antibodies to either MERS-CoV or SARS-CoV-1^76,77^. Because children experience less severe disease during SARS-CoV-1 infection than adults, it was hypothesized that childhood vaccination with non-CoVs provided cross-protection against SARS-CoV-1. However, binding and neutralizing antibodies and T cell responses induced by routine childhood vaccinations (AMPV, BCG, DPT, HBV, HIB, JEV, MMRV [and MV, RV], OPV, PI, SV, VV [varicella vaccine]) did not cross-react with SARS-CoV-1 in experimentally inoculated mice^78^.

#### Emerging HCoVs can induce cross-reactive binding antibodies toward endemic and other emerging HCoVs

SARS patients often experience a >4-fold rise in antibody to HCoV-229E, HCoV-NL63, and/or HCoV-OC43 in paired acute/convalescent samples (n=12/20)^39^. In one study, the number of SARS patients who experienced a >4-fold rise in binding antibodies was greater to HCoV-OC43 (n=10/11) than HCoV-229E (n=5/11)^73^. Some SARS patients also showed a rise in antibodies to HCoV-229E and HCoV-OC43 N protein^73^ and HCoV-NL63^79^. Among SARS-CoV-1 patients (n=28), 60% had detectable IFA titers to MERS-CoV and 25% had anti-MERS-CoV neutralizing antibodies. A subset with available paired samples experienced seroconversion to HCoV-OC43, but limited seroconversion to Alpha-CoVs. In the same study, animal handlers at a wildlife market in Guangzhou (n=94) with low-level prevalence of antibodies to SARS-CoV-1 (13.8% by IFA, 4.3% by NT) had detectable antibodies toward MERS-CoV (2.2% by IF)^76^. A follow-up study found that the cross-reactivity between SARS-CoV-1 and MERS-CoV was unlikely to be due to similarity in the receptor-binding domain (RBD), as monoclonal antibodies (mAbs) raised to SARS-CoV-1 RBD did not bind MERS-CoV RBD or neutralize MERS-CoV even at high concentrations^80^. MERS-CoV may produce less cross-reactivity against SARS-CoV-1. A subset of slaughterhouse workers in Saudi Arabia had antibodies to MERS-CoV by IFA as well as to pandemic HCoVs, but none had reactivity to SARS-CoV-1 spike protein. MERS-CoV patients were observed to have low-level cross-reactivity to SARS-CoV-1^77^ and in one study HCoV-HKU1 N protein^79^. Because of this cross-reactivity, researchers have developed diagnostic assays with truncated SARS-CoV-1 S, N, and M proteins^81–84^. See Figure 5 for associations between homologous and heterologous titers.

**Figure 5:**
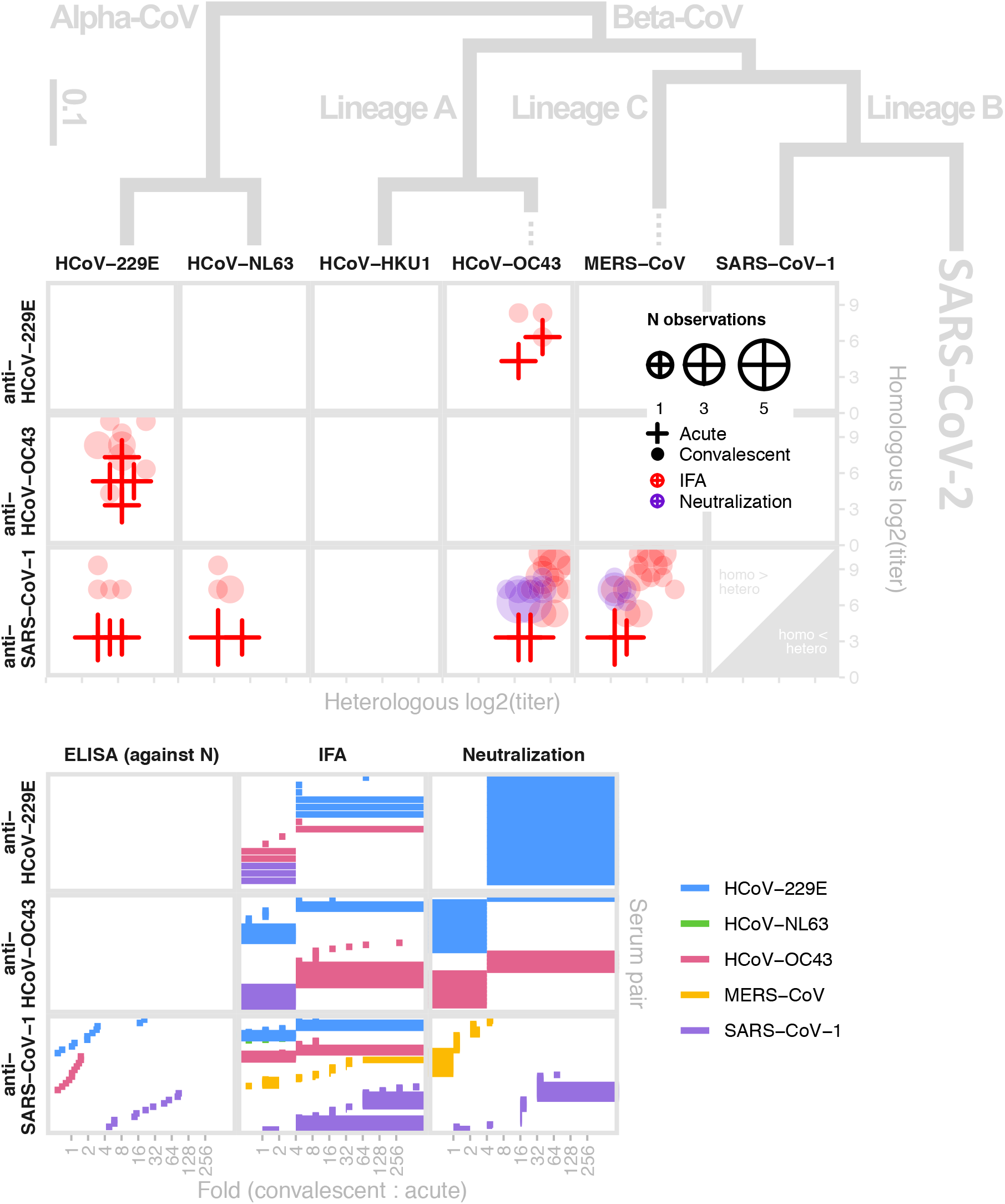
Antigenic and phylogenetic relationships among human coronaviruses. (Top) Reactivity of antiserum (x-axis) taken from individuals with confirmed infections of each human coronavirus against a panel of human coronaviruses (columns) shown in relation to their phylogeny as measured by IFA (red) and neutralization (purple); convalescent (circles) and acute sera (crosses); size of plot characters denotes the number of observations. The y-axis provides titers to the homologous strain the sera (rows) for comparison. (Bottom) Bracket of reported convalescent-to-acute titer fold-rise upon infection. Data extracted from 7 studies^39,65,69,72,73,139,140^. Phylogeny modified from ^141^.

#### There is some evidence for antigenic evolution in the receptor binding domain of emerging CoVs

A study of SARS-CoV-1 strains from the zoonotic phase (palm civets and bats) to early and late in the SARS epidemic revealed that some escaped neutralization by mAbs targeting the spike RBD of SARS-CoV-1^85–87^. Sera from BALB/c mice immunized with full length S protein from civet strains were ineffective against human SARS-CoV-1 and vice versa^88^. Despite significant cross-reactions between mAbs against conformational epitopes of RBD with multiple mutational differences, single mutations were shown to disrupt neutralizability^89^. In contrast, mAbs targeting regions critical for fusion and entry on the S2 protein are immunogenic and can broadly neutralize SARS-CoV-1 strains^59,90^. A recent study showed reduced binding of mAbs from SARS patients to the RBD of SARS-CoV-2, especially those that blocked binding to the ACE2 receptor. The only mAb potently bound to SARS-CoV-2 RBD protein did not compete with the RBD for binding to the ACE2 receptor, suggesting it bound a different, conserved site on the protein^91^. Similar studies were conducted to study changes in MERS-CoV. Five recombinant RBD proteins were constructed with mutations detected from MERS-CoV strains isolated in humans (2012-2015) and camels^92^. These RBDs maintained functionality and induced potent neutralizing antibodies. When residues in their receptor binding motifs were mutated to evade neutralization, cross-reactivity persisted but binding affinity to DPP-4 (the main receptor for MERS-CoV) was lost, suggesting limited antigenic escape for MERS-CoV. A study of MERS-CoV isolates with distinct amino acid differences in the S and other replication proteins found differences in replication kinetics, but it is not clear that these were attributable to differences in S^93^.

### Immunopathogenesis

In the initial review, 38 papers were identified as related to immunopathogenesis. Of these, we found 25 sufficiently relevant to review in Table S6, and 16 of those are detailed below and summarized in Figure 6.

**Figure 6:**
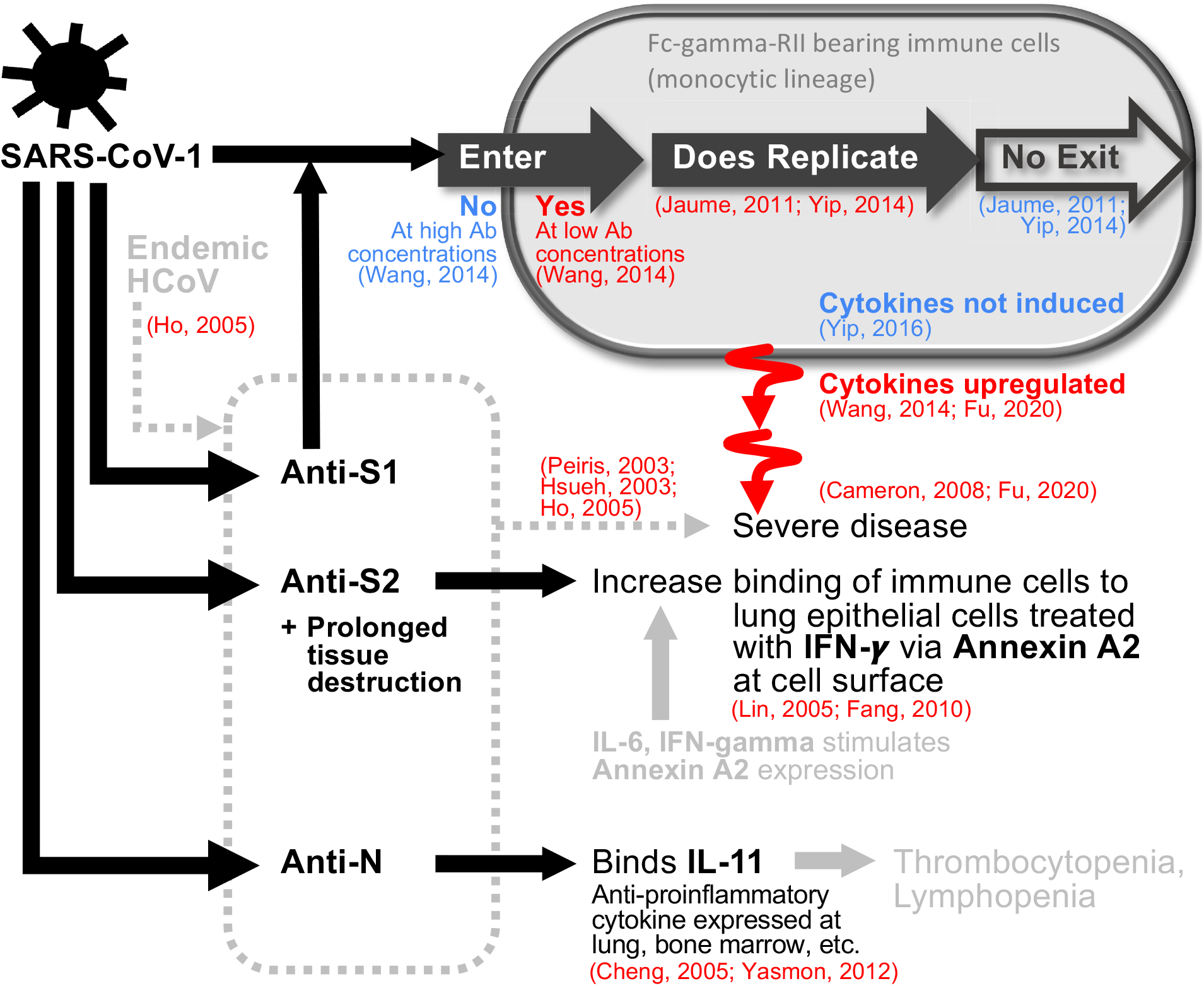
A diagrammatic summary of evidence supporting (red) and contradicting (blue) the contributions of antibodies to the pathogenesis of SARS-CoV-1. Anti-S1 antibodies triggered upon infection may facilitate entry into immune cells at later stages of the infection if concentration is low. Replication happens but no virus is released. Consequential induction of cytokines are inconclusive, but if they occur, they are associated with severe disease. Roles of anti-S2 and anti-N antibodies are supported by binding observations. Items shown in grey are weaker evidence given as speculations in the literature.

#### Antibody-dependent enhancement (*ADE*) *may be possible*

Multiple studies have suggested the possibility that pre-existing antibodies increase disease by facilitating viral entry into cells. This mechanism, a long hypothesized explanation for severe dengue infections, is called antibody-dependent enhancement (ADE). This has also been hypothesized as relevant to coronaviruses specifically in patients who seroconverted early in SARS-CoV-1 infection^25,94,95^. Reasons why some individuals can mount responses more quickly than others are unknown, but the correlation between greater age and early response with higher titers suggest priming effects from existing antibodies against endemic strains^95^. Authors from another study pointed toward enhancement from antibodies within the episode^17^. Nasopharyngeal viral load increased in the first week and declined thereafter but clinical worsening was seen in many of the patients at week two, with virus shedding in stool and urine observed towards the end^25,94^. Many manifested with new-appearing lesions as original lesions improved. The timing of the appearance of new lesions correlated with IgG seroconversion, suggesting that pathology post week one was driven by the immune response rather than by uncontrolled viral replication. Others have argued that ADE due to neutralizing antibodies is unlikely as treatment of SARS-CoV-1 with convalescent serum did not result in adverse effects^95^.

Observation of increased risk of disease in infants with mid-levels of maternal antibody to dengue (compared to high or low levels of maternal antibody) is important evidence supporting the role of antibody in pathogenesis for dengue^96^. A similar mechanism has been identified for seropositive and seronegative kittens infected with feline infectious peritonitis virus, a coronavirus that causes fatal disease in cats^97^. One study characterizing endemic HCoV in infants observed the highest lower respiratory tract infection burden (7.8%) at 6-23 months of age (1.5% at <6 months, close to none at 2-5 years) while upper respiratory tract infection burden was close to uniform^98^. The heightened disease burden after maternal immunity waning to medium levels in the lower tract, and the absence of such observation in the upper tract where antibodies do not circulate, subtly supports the possibility of ADE in HCoVs.

#### Suggestive enhancement mechanisms from in vitro experiments

Controlled *in vitro* experiments have explored the possible enhancing action of antibodies for HCoV infections. A series of studies by Yip et al. demonstrated that opsonization of anti-spike antibodies allowed SARS-CoV-1 to enter non-ACE2 expressing immune cells that bear Fc-*γ*-RII (CD32)^99,100^. Though replication is observed after entry, the virus does not exit nor alter the expression of proinflammatory immune mediators (CCL2/MCP-1, CCL3/MIP-1*α*, CXCL10/IP-10, TNF-*α*) and apoptosis inducing ligands (FasL)^101^. This contrasts with effects in nonhuman primate studies where endocytosis into macrophages stimulated inflammation, which in turn drove severe lung injuries^102^. A study by another group in human promonocyte cell line (HL-CZ), which expresses both ACE2 and Fc-*γ*-RII, demonstrated increased infectivity and virus-induced apoptosis when anti-SARS-CoV-1 sera from patients were added at 100-to 2000-fold dilutions, while at higher concentrations neutralization occurred^103^. Upon infection, TNF-*α*, IL-4 and IL-6 expressions heightened, while IL-3 and IL-1*β* only appeared in trace amounts. The difference could have resulted from the cell line differences or the set of mediators assessed^104^. Regarding regions of the spike protein that may induce antibodies with enhancement effects, both anti-S1a and anti-S1b mAbs showed mild to moderate effects^103^. Only one particular anti-S1b clone showed neutralization. No effect was seen for anti-N mAbs. There is limited evidence that these mechanisms are causal to inflammatory gene expression differences among patients with differing severities^105^.

#### Direct damage caused by antibodies

While the link between antibody presence and enhanced severity via infection of immune cells remains unclear, some have suspected a role for auto-reactive responses in increased severity of illness. Prolonged tissue destruction can increase presentation of host proteins to T- or B-cells and result in an adaptive response against self, i.e. epitope spreading^52^. Anti-S2 IgG antibodies targeting uninfected lung epithelial cells (A549) were detected in SARS-CoV-1 patients after twenty days post-symptom onset^106^, a reactivity not seen in serum from healthy individuals and non-SARS-CoV-1 pneumonia patients. Complement inactivation only showed cytotoxic effect when IgG was present/unbounded. Presence of anti-S2 antibodies also increased the binding of immune cells (PBMCs) to A549 cells treated with IFN-*γ*, replicating the conditions under which a cytokine storm would be observed. A separate study demonstrated colocalization of anti-S2 Abs collected from serum of SARS patients ≥50 days post-fever onset with annexin A2 and immunoprecipitated annexin A2 on A549 cell surfaces. Elevated expression of annexin A2 on the surface can be stimulated by IL-6 and IFN-*γ*, both cytokines induced by SARS-CoV-1, which in turn increases binding of anti-S2 Abs to the cell. However, its pathogenic role was not explored^107^. Alternatively, similarity between viral and host epitopes (molecular mimicry) can generate cross-reactive antibodies. In mice^108^ and humans^109^, there is evidence of anti-N antibodies that cross react with IL-11, an anti-inflammatory cytokine expressed in many tissues, including lung and bone marrow. The authors suggest high anti-N antibodies induced relatively early during infection may be involved in the thrombocytopenia and lymphopenia observed early in SARS-CoV-1 infection^108^.

### Population seroprevalence

From the paper review, 68 papers were classified as having data on age and seroprevalence or seroincidence, of which 20 studies were confirmed on further review, and 14 had digitizable data. The age range across studies was 0 to ≥65, while the sample size of studies ranged from 69-19,974 (82-6,400 for studies of endemic HCoVs). Table S7 and Supplementary Data File S3 contain details of the studies.

#### Seroprevalence to endemic coronaviruses increases rapidly during childhood

For endemic coronaviruses, seroprevalence rises sharply in childhood, with little to no change in seroprevalence by age among adults. While exact dynamics from 6 months to 20 years vary between studies, the general trend remains consistent. Figure 7 displays HCoV age-seroprevalence curves for the 7 papers with digitizable data on seroprevalence by age, with the panels representing the four major endemic strains^74,110–115^. Trends from papers not displayed in the figure are largely the same^68,70,116,117^. Two studies show a marked decline in seroprevalence with age above 40^68,111^.

**Figure 7:**
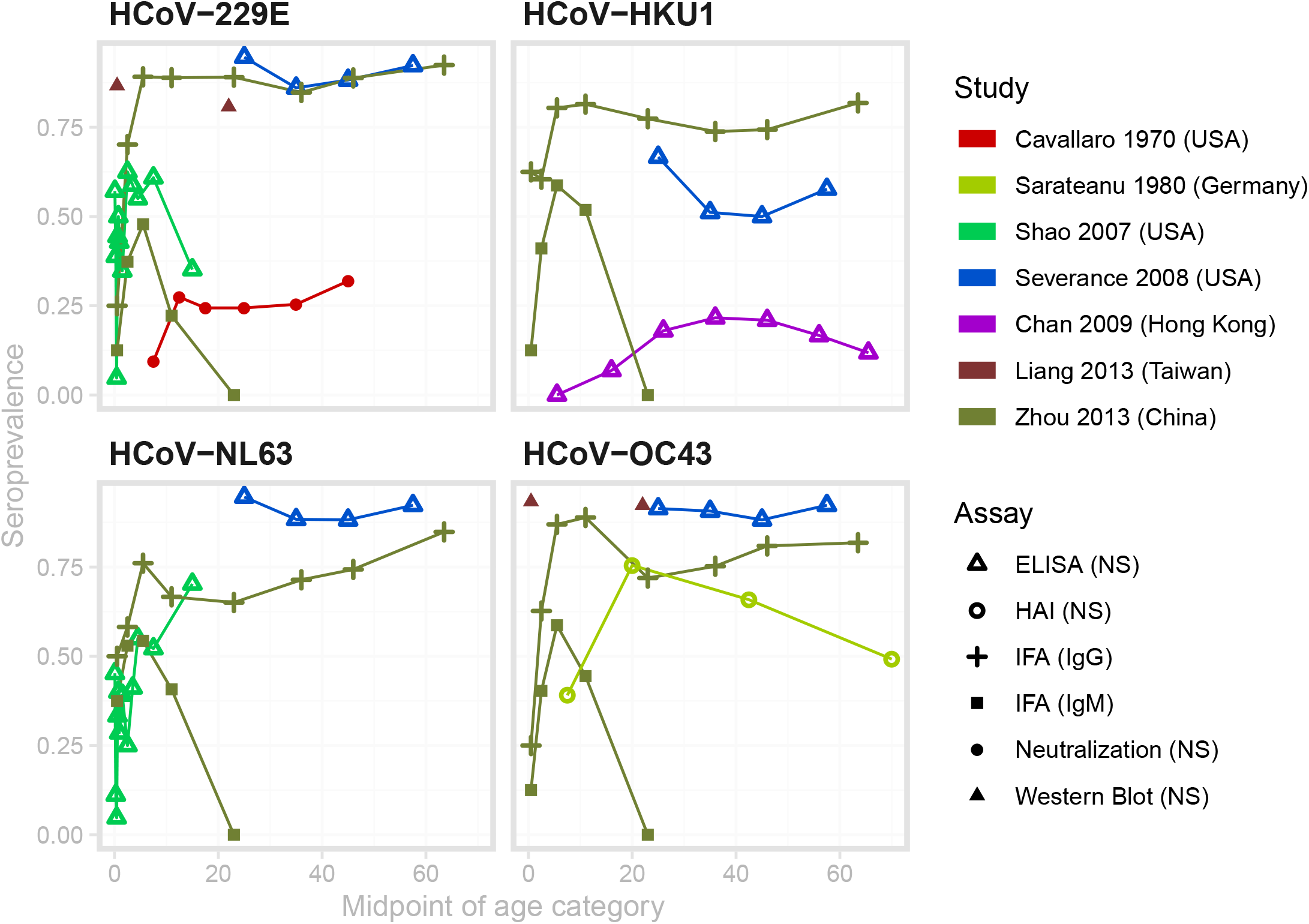
Age-seroprevalence curves for the studies with appropriate, digitizable data on endemic HCoV. The color denotes the study, and the point type denotes the assay and antibody measured. Data from Sarateanu et al is averaged over two serosurveys conducted in 1975 and 1976. Data from Liang et al. were measured in cord blood samples and adults aged 18-25, and do not represent the change in seroprevalence across childhood. Therefore we have not connected these two points with a line.

#### Seroincidence of endemic coronaviruses occurs in all age groups, with no clear trend

Available data sets are more sparse for incidence, and the patterns are inconsistent between studies^110,118–120^. Figure 8 displays age-incidence curves for the 4 papers with digitizable data on incidence by age, with each panel representing the two strains included in these studies (HCoV-229E and HCoV-OC43). There was an increasing trend in seroincidence with age during an outbreak for HCoV-229E in Tecumseh, Michigan^110^, but a similar analysis of HCoV-OC43 seroincidence in the same outbreak showed no such increase^118^. Longitudinal follow-up of 10 families in Seattle found a lower rate of seroconversions among adults compared to children^119^. A comparison of a cohort of 21-40 year-olds and a cohort of ≥65 year-olds found no clear difference in incidence between the two age groups, as measured by seroconversion or PCR-confirmed, HCoV-associated respiratory illness^120^. Finally, observational studies have shown evidence of coronavirus-associated respiratory illnesses in elderly populations^121–123^ and across all ages^124^.

**Figure 8:**
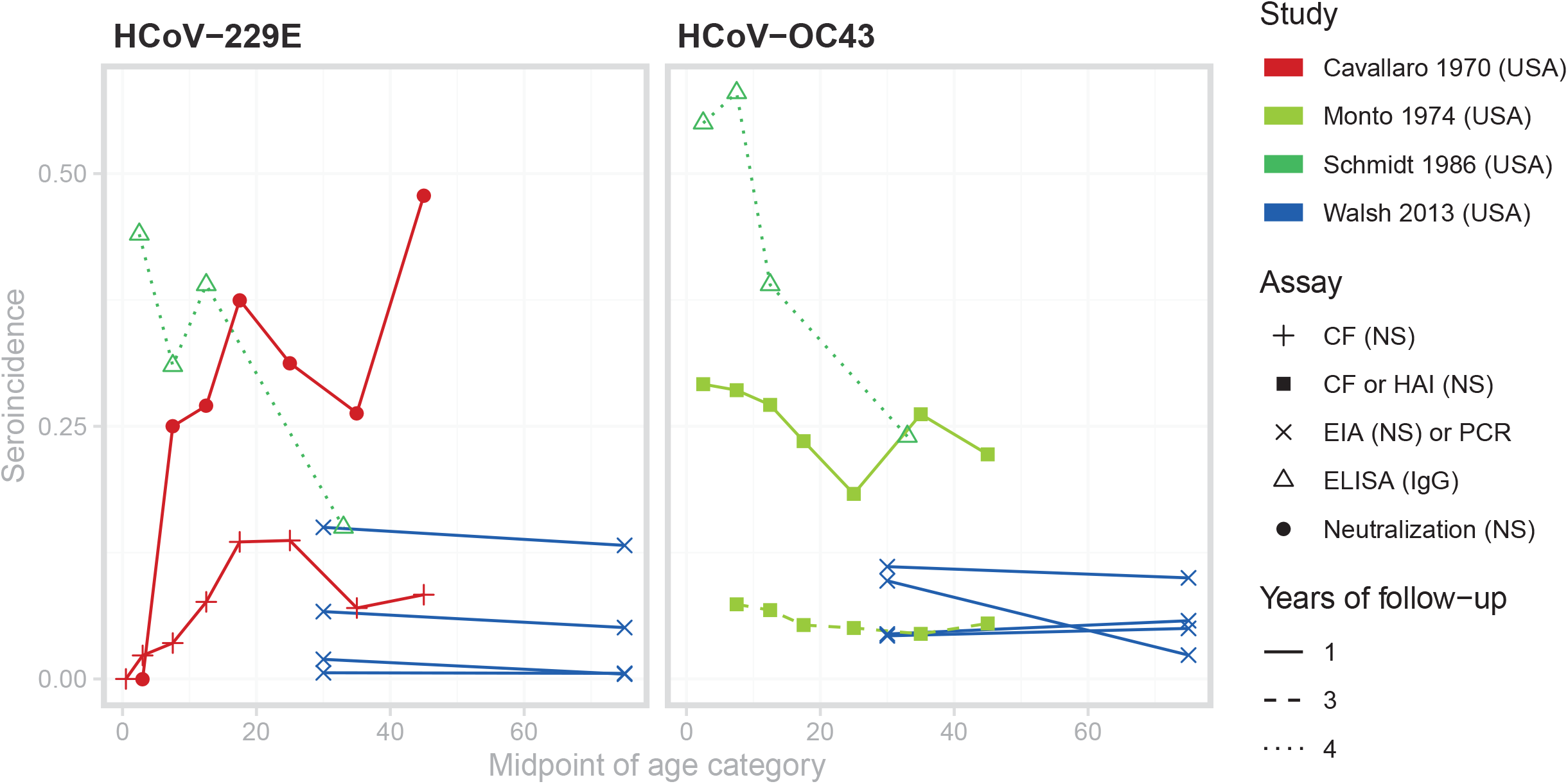
Age-incidence curves for studies with appropriate, digitizable data on endemic HCoV. The color denotes the study, the point type denotes the assay and antibody measured, and the line type represents the number of years between successive serosample

The force of infection of endemic coronavirus strains is high, and the age at first infection low, but variable across studies. Simple catalytic models fit to the digitized data predict median total force of infection across studies of 0.21 (95% CI 0.09, 0.40) (see Table S8 for estimates by study), corresponding to an average age at first infection with any strain of 4.8 years (95% CI 2.5, 11.2). A cohort of 25 infants followed from birth for an average of two years experienced annual strain-specific incidence rates from 0.12-0.70^70,71^. Zhou et al.’s serosurvey measured IgG and IgM separately, and showed very different patterns with age^115^ (Figure 7). While IgG seroprevalence rose to high levels by age 10 and remained high into the adult population, IgM seroprevalence declined to zero for all individuals aged 14 and older. The authors interpreted this as evidence that first infections occurred before the age of 14 for all strains.

#### Data are incompatible with complete, lifelong homologous immunity to endemic coronaviruses

The rapid rise in seroprevalence with age observed in the literature suggests a force of infection (and thus proportion seropositive to all serotypes at older ages) high enough to preclude significant rates of coronavirus infection among older adults if each serotype confers lifelong homologous immunity (Figure 9, green dotted line). On the other hand, short-term, complete immunity, lifelong partial immunity, and the existence of multiple genotypes within a single serotype with limited cross-immunity could explain the age-seroprevalence and age-incidence curves seen in the literature (Figure 9).

**Figure 9:**
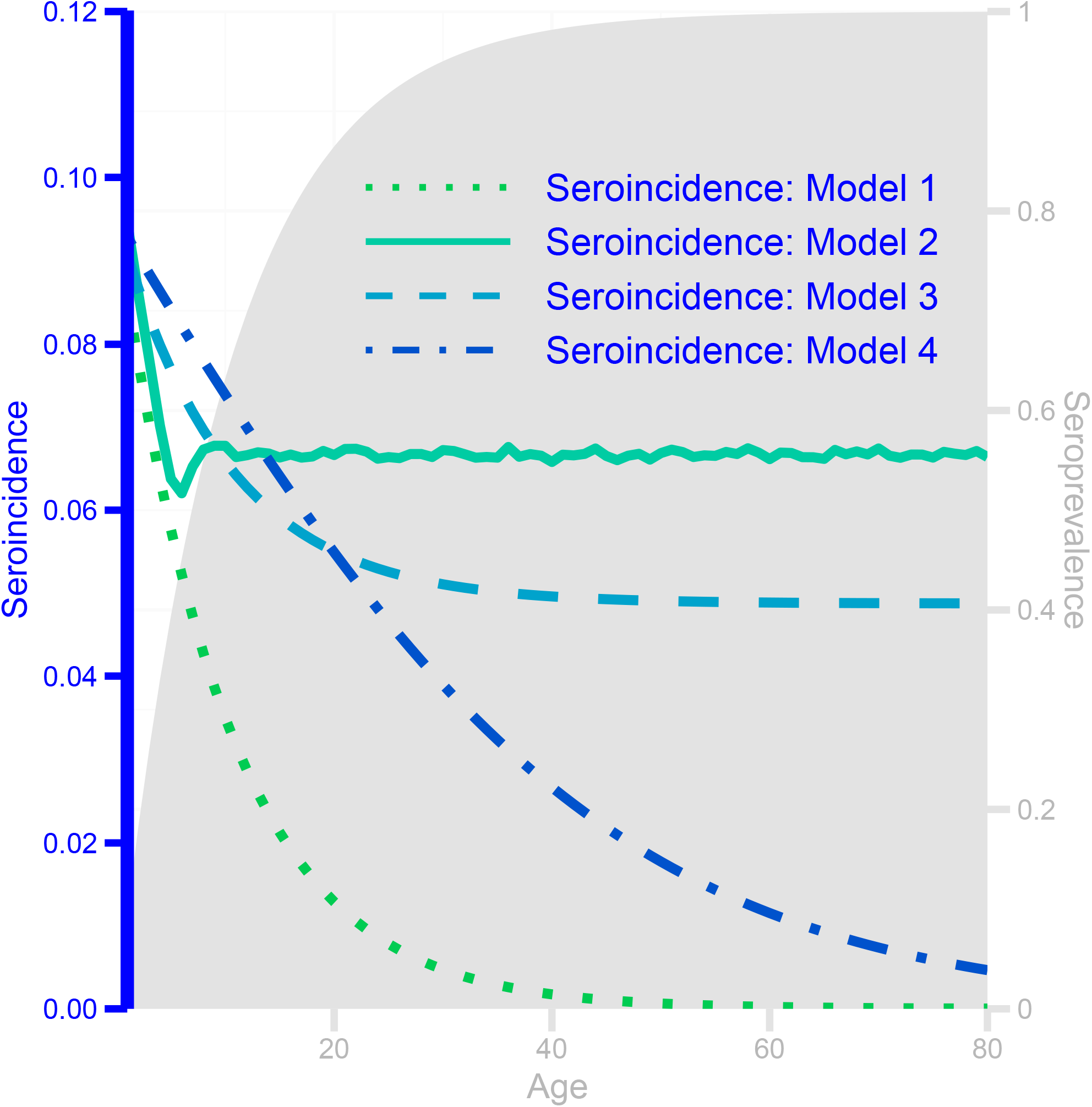
Seroprevalence (shaded area, scale indicated by right y-axis) and seroincidence (lines, scale indicated by left y-axis) curves by age for a single strain for four hypothetical models of coronavirus immunity: complete homologous immunity (Model 1), complete homologous immunity for 5 years, then reversion to full susceptibility (Model 2), partial homologous immunity for life after the first infection (Model 3), and 4 genotypes within a strain, conferring lifelong genotype-specific immunity but no within-strain cross-immunity (Model 4).

#### Further observations from review

The review gave rise to several observations that were either consistent or conflicting across papers. Studies that enrolled children less than 6 months detected loss of maternal antibodies, representing a preliminary line of defense for newborn children^70,71,74,117^. Most studies showed no discernible difference in age-seroprevalence trends or in overall seroprevalence by strain. Gao et al.^68^ found that seroprevalence of HCoV-229E and HCoV-HKU1 was significantly lower than seroprevalence of HCoV-OC43 and HCoV-NL63. In addition, Chan et al.^114^ found seroprevalence to HCoV-HKU1 to be low (21.6% in 31-40 year olds) in Hong Kong, and stated that this was expected from the low rates of HCoV-HKU1 among respiratory illnesses. Finally, most studies measured the presence of binding antibodies in the blood. Of the studies in Figure 7, only Cavallaro and Monto^110^ (red) measured seropositivity using a neutralization assay. That the seroprevalence is markedly lower in that study compared to the others could indicate lower test sensitivity compared to more modern tests, or of the lack of correlation between neutralizing and binding antibodies against endemic coronaviruses.

Regarding the novel coronaviruses, serosurveys of SARS-CoV-1 confirmed that the rate of asymptomatic or subclinical infection was very low^125^. Asymptomatic and subclinical rates of MERS-CoV are generally higher, but available serosurveys lack the power to draw strong inference about age trends. A large serosurvey conducted across Saudi Arabia found that the age of seropositive individuals was significantly lower than the age of clinical cases^126^, while another conducted across multiple countries in Africa and Asia found no trend in seroprevalence with age^127^. Studies of risk factors within camel workers have either not addressed age as a risk factor^128,129^ or not found an association^130^.

## Discussion

### Key findings

We have presented a broad, comprehensive review of multiple aspects of the literature on antibody immunity to coronaviruses. We identified a number of key findings. The median time to detection was similar across different antibodies for SARS-CoV-1 (12 days; IQR 8-15.2 days) and SARS-CoV-2 (11 days; IQR 7.25-14 days), but longer for MERS-CoV (16 days; IQR 13-19 days). Most long-term studies found that IgG waned over time (typically detectable up to at least a year) while others found detectable levels of IgG three years post symptoms onset. Antibody kinetics varied across the severity gradient with longer durations of detectable antibody associated with more severe symptoms. Human challenge studies with HCoV indicate that serum and mucosal immune responses (serum IgG, IgA, neutralizing titer, mucosal IgA) provide possible correlates of protection from infection and disease. However, repeat human challenge experiments with single HCoV suggest individuals can be infected with the same HCoV one year after first challenge, but with possible lower severity. There is cross-reactivity within but minimal reactivity between Alpha- and Beta-CoVs. While endemic HCoVs rarely induce cross-reactive antibodies against emerging HCoVs, SARS-CoV-1 and MERS-CoV stimulate antibodies induced by prior HCoV infections. Multiple mechanisms for immunopathology have been suggested but no strong causal evidence exists and the extent to which the presence of antibodies affects human disease severity is not known. Seroprevalence with the four major endemic HCoV strains rose rapidly during childhood and remained high in adults. The median age at first infection with any strain was 4.8 years (95% CI 2.5, 11.2). There was no clear trend in seroincidence with age, and many studies have demonstrated incidence of coronavirus infections in elderly populations. These results suggest a measurable impact of immunity to coronaviruses on future risk, but this protection may be transient.

### Limitations of systematic review

We have suggested a set of search terms to identify relevant work, and others may expand on this search. Due to the speed of new research being produced, we limited our systematic review of SARS-CoV-2 papers to before March 20, 2020. Further evidence on immune responses to SARS-CoV-2 is likely contained in work that has since been published. We excluded from the review animal studies and studies of animal CoVs to remain human-focused, but this literature is likely relevant to some areas of the review. We limited the scope of the review to antibody-mediated immunity, meaning that understanding of some areas of the review may be incomplete. Finally, digitized data could be affected by publication bias or other selection bias, meaning that it might not be representative of data from all studies. The aim of the pooled analyses was to summarize the array of studies rather than formally explore hypotheses, and as a result the models used are simple. For example, in estimating force of infection from age-stratified seroprevalence data, we did not consider other features of a model such as assay sensitivity, time- and age-varying force of infections, seroreversion, and cross-reactivity between strains, that might better explain patterns seen in the data.

### Implications for SARS-CoV-2 pandemic responses and future outlooks

#### Serological assay development

There is a **need for development of serological assays with high sensitivity for screening and sufficient specificity to exclude individuals from unneeded interventions**. Evidence from previous emerging CoVs suggests that false negativity is likely to result from waning of antibody levels. As most studies on antibody kinetics are on symptomatic patients, the kinetics in subclinical infections, a significant proportion of SARS-CoV-2 infections^131^, remains a key gap in the literature. Indications that less severe illnesses are associated with reduced antibody responses suggests that mild cases may pose challenges to serological assays.

On the other end, the proposed policy of **“immunity passports”** to allow individuals presumed immune to return to work once social distancing measures are relaxed **requires highly specific serological tests** to mitigate adverse outcomes. Evidence from past emerging HCoVs suggests low false positivity from cross-reaction with endemic HCoVs. However, antibody titers do not necessarily translate to immunity. Challenge studies indicate multiple candidates that may serve as correlates of protection including serum and mucosal measures; however, these will need specific evaluation for SARS-CoV-2. The knowledge gap in correlates of protection and their durability must be filled before immunity passports are safe for general use.

#### Therapeutics, vaccines, and ADE

Pharmaceutical interventions to protect susceptible individuals or improve outcomes among sick individuals will first need to be tested in randomized trials. Vaccines with various antigens are currently under development^132^ and polyclonal antibodies from SARS-CoV-2-recovered patients have been used in some cases for treatment^133^. Though the evidence for immunopathogenesis remains limited^134^, it will be important to design rigorous trials that can provide valid evidence of positive effect but also detect negative effects if they occur with associated adaptive stopping criteria. Such concerns might extend beyond vaccine development if negative effects are subtle and not or weakly detectable in Phase III studies. In the event of mass vaccination following successful efficacy studies, surveillance designed to detect waning immunity and adverse events due to vaccine-mediated immunopathogenesis is critical.

#### Long-term dynamics of SARS-CoV-2 and concluding remarks

Key questions for the long-term dynamics of SARS-CoV-2 include whether it is likely to be eradicated, whether widespread immunity will lead to transient decreases in incidence and how long it will take for another large outbreak to occur. Modeling studies will be crucial in answering these questions, but they will rely on an understanding of immune responses to SARS-CoV-2 and interactions with other coronaviruses to make relevant predictions. Analysis of datasets, specifically of serosurveys, needs to account for the kinetics of waning immunity and effects of imperfect assays to draw meaningful inferences.

Finally, we note that the best evidence for immune responses to SARS-CoV-2 will come from studies of the virus itself. Such studies are being performed and reported with unprecedented speed, but in the absence of firm evidence many are turning to the existing coronaviruses as a model. To that end we have produced this review to aid researchers in understanding the scope of the evidence for immune responses to these coronaviruses.

## Data Availability

We have made all data related to our systematic review and pooled analyses available at the following archive:
https://doi.org/10.5281/zenodo.3751566

## Acknowledgements

This research was supported [in part] by the Intramural Research Program of the National Human Genome Research Institute, National Institutes of Health. DATC was supported by the US National Institutes of Health award number R01-AI114703-01.APW is funded by a Career Award at the Scientific Interface from the Burroughs Wellcome Fund and by the National Library of Medicine of the National Institutes of Health under award number DP2LM013102. IRB was supported by the John A Watson Faculty Scholar fellowship. We thank Celeste Dale and Maxwell Hogshead for assistance in data digitization.

## Supplementary Figure Titles

Figure S1: **Cumulative proportion of patients that seroconverted**.

Figure S2: **Antibody time series reported in studies in units of optical density (OD)**. Figure S3: **Antibody time series reported in studies in units of titers**.

## Supplementary Table Titles

Table S1: **Summary of serological Assays**

Table S2: **Summary of studies on the kinetics of antibody immunity after infection, and for the association of antibody responses with disease severity**.

Table S3: **Severity ratings used in the studies, and the corresponding standardisations**.

Table S4: **Summary of studies on correlates of antibody immunity and protection against CoVs infection**.

Table S5: **Summary of studies on antigenic diversity, cross-reactivity**.

Table S6: **Summary of immunopathogenesis**.

Table S7: **Summary of studies on population-level seroprevalence of CoV**.

Table S8: **Estimated annual force of infection from digitized age-seroprevalence data for endemic HCoVs**.

## Supplementary Data Titles

Supplementary Data S1**: Data digitized on antibody kinetics and association of antibody responses with clinical severity**.

Supplementary Data S2: **Data digitized on cross-reactivity and antigenic diversity**.

Supplementary Data S3: **Data digitized on population seroprevalence**.

